# Validation and clinical evaluation of a SARS-CoV-2 Surrogate Virus Neutralization Test (sVNT)

**DOI:** 10.1101/2020.09.21.20191288

**Authors:** Benjamin Meyer, Johan Reimerink, Giulia Torriani, Fion Brouwer, Gert-Jan Godeke, Sabine Yerly, Marieke Hoogerwerf, Nicolas Vuilleumier, Laurent Kaiser, Isabella Eckerle, Chantal Reusken

## Abstract

To understand SARS-CoV-2 immunity after natural infection or vaccination, functional assays such as virus neutralizing assays are needed. So far, assays to determine SARS-CoV-2 neutralizing antibodies rely on cell-culture based infection assays either using wild type SARS-CoV-2 or pseudotyped viruses. Such assays are labour-intensive, require appropriate biosafety facilities and are difficult to standardize.

Recently, a new surrogate virus neutralisation assay (sVNT) was described that uses the principle of an ELISA to measure the neutralization capacity of anti-SARS-CoV-2 antibodies directed against the receptor binding domain.

Here, we performed an independent evaluation of the robustness, specificity and sensitivity on an extensive panel of sera from 269 PCR-confirmed COVID-19 cases and 259 unmatched samples collected before 2020 and compared it to cell-based neutralization assays. We found a high specificity of 99.2 (95%CI: 96.9-99.9) and overall sensitivity of 80.3 (95%CI: 74.9-84.8) for the sVNT. Clinical sensitivity increased between early (< 14 days post symptom onset or post diagnosis, dpos/dpd) and late sera (>14 dpos/dpd) from 75.0 (64.7-83.2) to 83.1 (76.5-88.1). Also, higher severity was associated with an increase in clinical sensitivity. Upon comparison with cell-based neutralisation assays we determined an analytical sensitivity of 74.3 (56.4-86.9) and 98.2 (89.4-99.9) for titres ≥10 to < 40 and ≥40 to < 160, respectively. Only samples with a titre ≥160 were always positive in the sVNT.

In conclusion, the sVNT can be used as an additional assay to determine the immune status of COVID-19 infected of vaccinated individuals but its value needs to be assessed for the specific context of use.

## Introduction

In 2020, the world is facing an unprecedented global health crisis through the emergence of the novel coronavirus severe acute respiratory syndrome coronavirus 2 (SARS-CoV-2), the causative agent of the disease COVID-19. The pandemic spread of this virus immediately raised a demand for serological assays to support clinical and public health management, e.g. to determine a recent or past infection, to assess the level of (sub) population exposure and, to investigate different types of immune response and levels of potential immunity against re-infection. Seven months into the outbreak a plethora of serological assays is available^1^ that allows the routine detection of several classes of antibodies, i.e. IgM, IgG and IgA^2–5^. However, to understand immunity after natural infection or vaccination, a functional analysis of the elicited antibody responses, such as avidity for the most immunogenic viral antigens and virus neutralizing activity, is of utmost importance^6^.

So far, assays to determine SARS-CoV-2 neutralizing capability of antibodies rely on handling of wild type or pseudotyped viruses and use cell-culture based infection as a read-out. This requires a biosafety level (BSL) 3 laboratory for wild type SARS-CoV-2, or a BSL-2 laboratory for pseudotyped viruses such as vesicular stomatitis virus (VSV) or lentivirus-based systems. These in-house assays are difficult to standardize across laboratories, especially in the absence of an international standard, as assay characteristics vary depending on culture conditions, virus strains and cell lines used. Furthermore, these assays are labour-intensive, require highly skilled personnel, have a low throughput and results are only available after several days. Recently, a first commercial assay has become available^7^ that indirectly and semi-quantitatively measures the neutralizing functionality of SARS-CoV-2 antibodies while overcoming the above limitations. During natural infection, SARS-CoV-2 binds to its cellular receptor, the angiotensin-converting enzyme 2 (ACE2), via the receptor binding domain (RBD) of the viral spike (S) glycoprotein, which is an essential step to establish infection of the cell^8,9^. The majority, but not all, of the neutralising antibodies are directed against the RBD leading to an inhibition of this interaction^6^. The assay detects SARS-CoV-2 antibodies that competitively inhibit the interaction between recombinant RBD-HRP fusion protein and recombinant ACE2 that is coated on 96-well plates. The assay is independent of the use of replicating or pseudotyped virus and cell cultures and uses the same format/set-up as Enzyme-Linked Immunosorbent Assays (ELISA), allowing for high-throughput, automation and fast turnaround times.

Here, we present an independent, two-centre evaluation of the robustness, specificity and sensitivity of a commercially available version of this novel functional immune-assay based on an extensive panel of sera from a) a heterogeneous cohort of PCR-confirmed COVID-19 patients, b) pre-outbreak syndromic patients with respiratory complaints including confirmed recent infections with the four common human coronaviruses (HCoV) and c) pre-outbreak population sera. The assay performance was evaluated against the conventional cell culture-based wildtype SARS-CoV-2 (Gold Standard method) and VSV-based pseudo-type neutralization assays to assess its value to measure levels of functional antibodies directed against SARS-CoV-2.

## Material and Methods

### Sample collection

RIVM: sera from common CoV cases and non-CoV respiratory cases were partially obtained from a previous study approved by the ethics committee of the National Institute of Public Health and the Environment (METC Noord-Holland, http://www.trialregister.nl; NTR3386 and 4818^10^ and partially from anonymized leftover serum from routine diagnostics for respiratory pathogens or SARS-CoV-2 for which ethical approval was waived by the ethics committees of Brabant and Utrecht (NW2020-31 and NL13529.041.06; 06/282). The current study was performed in accordance with the guidelines for sharing of patient data of observational scientific research in emergency situations as issued by the Commission on Codes of Conduct of the Federation of Dutch Medical Scientific Societies (https://www.federa.org/federa-english).

University of Geneva/HUG: Anonymized leftovers of serum and plasma samples were used for this analysis. Ethical approval for all samples used in this study was waived by the local ethics committee of the HUG that approves usage of leftover of patient samples collected for diagnostic purposes in accordance with our institutional and national regulations.

The study included blood samples from 269 real-time (RT)-PCR confirmed COVID-19 cases (sensitivity panel) and 259 unmatched samples collected before 2020 (specificity panel). We used days post onset of symptoms (dpos) in cases where the onset was known or days post PCR diagnosis (dpd) if the onset was unknown. Samples were stored at −20°C and thawed immediately before the assay was performed. The specificity panel included sera collected before 2020 from healthy blood donors (n=100), patients with other respiratory diseases including (i) a two months earlier PCR-confirmed common HCoV infection, HCoV-229E (n=12), HCoV-NL63 (n=10), HCoV-HKU1 (n=6) or HCoV-OC43 (n=10)^10^, (ii) patients with a recent PCR-confirmed non-CoV respiratory infection (n=14) i.e. Influenza A virus (n=3), human metapneumovirus (HMPV) (n=4), respiratory syncytial virus (RSV) B (n=1), RSV A + HMPV (n=1), hemophilus influenza (n =1), mycoplasma pneumonia (n=1) and rhinovirus (n=3). Further it included patients with respiratory complaints that tested negative for a suspected Bordetella pertussis infection (n=16), patients with an acute cytomegalovirus (n = 10) or acute Epstein-Barr virus (n=10) infection as well as adult (n=21) and child (n=50) patients who came for routine diagnostic purposes to the hospital (Table 1). The sensitivity panel included sera from 269 PCR-confirmed COVID-19 patients. Of these sera, 92 were taken before and 177 after 14 dpos/dpd. Severity of disease ranged from asymptomatic (n=3) to mild (non-hospitalized, n=92), severe (hospitalized, n=87) and ICU-admitted/deceased (hospitalized, n=54). For 33 patients the severity of disease was unknown (Table 1).

**Table 1:**
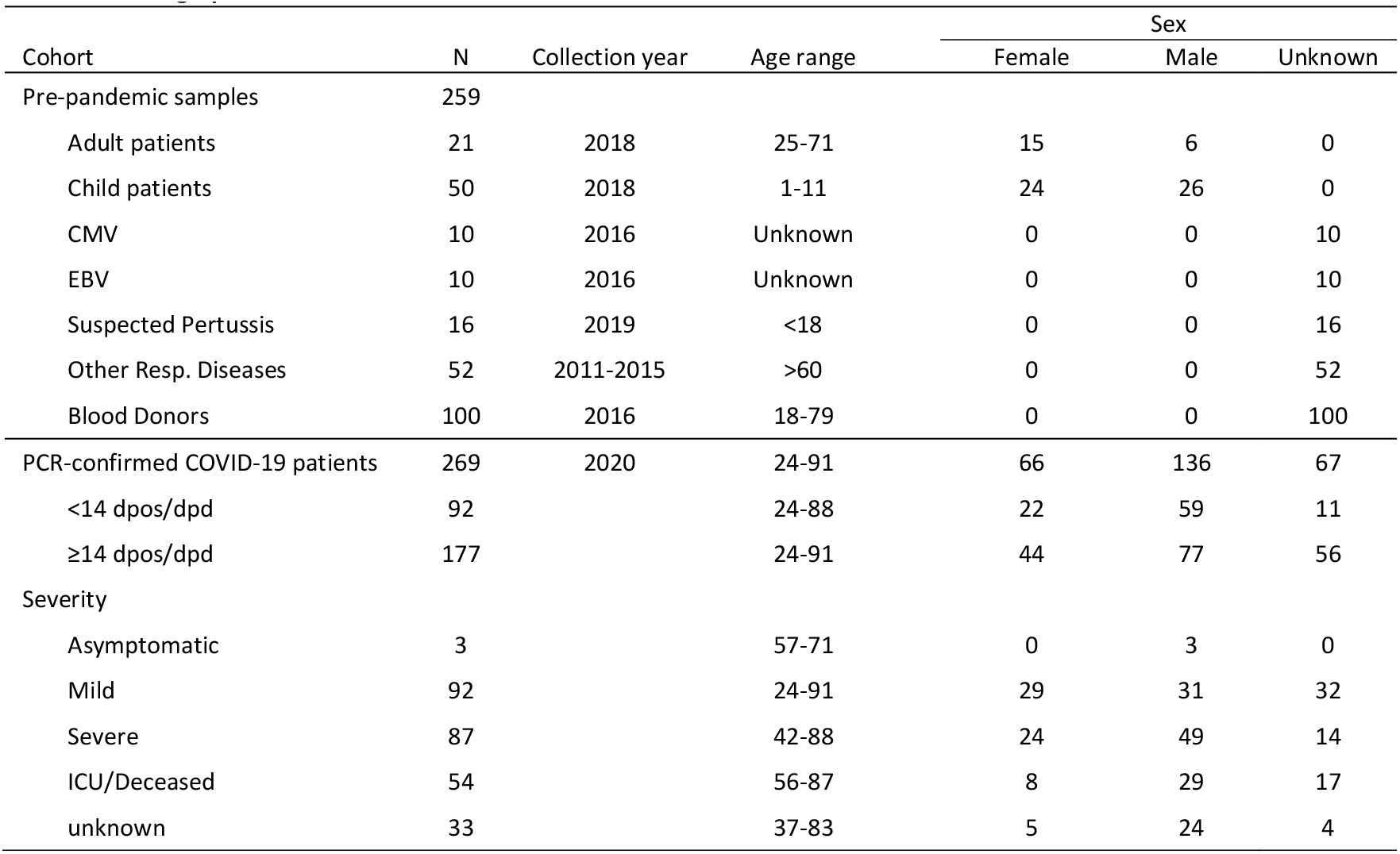
Demographics

### Surrogate SARS-CoV-2 Virus Neutralisation Test

The surrogate virus neutralisation test (sVNT) (GenScript cPass™ SARS-CoV-2 Neutralization Antibody Detection Kit, Genscript, The Netherlands) was performed according to the manufacturer’s instructions. Both laboratories used the same LOT number (20E012157). Briefly, serum samples as well as positive and negative assay controls were diluted 1:10 in sample dilution buffer and mixed with an equal volume of HRP-conjugated RBD. Controls were tested in duplicates and samples in singular. After a 30 minute incubation at 37 °C, 100 µl of this mixture was transferred to a 96-well plate coated with recombinant ACE2. After incubation at 37 °C for 15 minutes, the supernatant was removed and the plate was washed 4x using the provided wash buffer. 100 µl TMB substrate was added and incubated for 15 minutes at room temperature before the reaction was stopped by addition of 50 µl stop solution. Plates were read at 450 nm immediately afterwards. Percentage reduction (%reduction) for each sample was calculated by using the following formula:

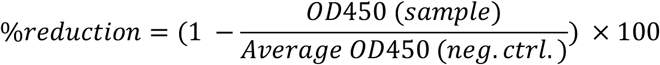

### VSV-based Pseudovirus Neutralisation Test (PNT50)

The VSV-based pseudovirus neutralisation test, was done as described previously^2^. Briefly, African green monkey (VeroE6) cells were seeded in 96-well plates at 2 × 10^4^ cells per well and grown into confluent monolayer overnight. Sera from patients were inactivated at 56°C for 30 minutes and diluted from 1:5 to 1:1280 in DMEM 2% fetal bovinve serum (FBS). VSV-based SARS-CoV-2 pseudotypes (generated according to Berger, Rentsch, and Zimmer^11^ and Torriani et al.^12^ expressing a 19 amino acids C-terminal truncated spike protein^13^ (NCBI Reference sequence: NC_045512.2) were diluted in DMEM 2% FBS in order to have MOI=0.01 per well and added on top of serum dilutions (final serum dilutions obtained were from 1:10 to 1:2560). The virus-serum mix was incubated at 37°C, for 2h. Vero E6 were then infected with 100µl of virus-serum mixtures. After incubation at 37°C for 1.5h, cells were washed once with 1X PBS and DMEM 10% FBS was added. After 16-20h of incubation at 37°C, 5% CO_2_ cells were fixed with 4%formaldehyde solution for 15min at 37°C and nuclei stained with 1µg/ml DAPI solution. GFP positive infected cells were counted with ImageXpress® Micro Widefield High Content Screening System (Molecular Devices) and data analyzed with MetaXpress 5.1.0.41 software.

### SARS-CoV-2 Virus Neutralisation Test

SARS-CoV-2 virus neutralization tests were performed exactly as described^14^. Two-fold serial dilutions (starting at 1:10) of heat-inactivated sera (30 min, 56°C) were incubated in duplicate with 100 TCID_50_ of SARS-CoV-2 strain HCoV-19/Netherlands/ZuidHolland_10004/2020 (EVAg cat.nr. 014V-03968) at 35°C, 5% CO_2_ for 1 h in 96-wells plates. Vero-E6 cells were added in a concentration of 2 × 10^4^ cells per well and incubated for three days at 35°C in an incubator with 5% CO_2_. The serum virus neutralization titre (VNT_50_) was defined as the reciprocal value of the sample dilution that showed a 50 % protection of virus growth. Samples with titres ≥ 10 were defined as SARS-CoV-2 seropositive.

### Wantai SARS-CoV-2 total antibody Assay

The Wantai SARS-CoV-2 total antibody ELISA (Beijing Wantai Biological Pharmacy Enterprise, Beijing, China; catalogue number WS1096) was performed exactly according to the manufacturer’s instructions^3^. This assay is a double-antigen sandwich ELISA using a recombinant RBD of SARS-CoV-2 as antigen. Optical density (OD) is measured at 450 nm and the antibody titre for each sample is calculated as the ratio of the reading of that sample to the reading of a calibrator (included in the kit): OD ratio.

### Statistical analysis

Statistical analysis was performed with GraphPad Prism version 8.4.3 using Mann-Whitney or Kruskal-Wallis test with Dunn’s multiple comparison test where appropriate. Linear regression was also performed using GraphPad Prism version 8.4.3. Calculation of sensitivity, specificity and 95% confidence intervals (95%CI) was done using the VassarStats statistical toolbox (http://www.vassarstats.net/). Results with p values <0.05 were considered significant.

## Results

In this study, we validated an ELISA-based surrogate SARS-CoV-2neutralisation assay (sVNT) using specificity and sensitivity panels as described above. Among the 259 samples in the specificity panel we identified two samples, one among the blood donors and one in the cohort with a recent non-CoV respiratory infection (HMPV infected) that were positive using the manufacturer recommended cut-off of 20% reduction (%reduction: 97.2 and 21.6, respectively). Both samples were tested in the VNT_50_ and did not show any SARS-CoV-2 neutralising activity. Considering an alternative cut-off of 30% reduction, as proposed in a recent publication by the manufacturer^7^, only the blood donor sample remained positive for blocking RBD binding to ACE2 (Figure 1A). This indicates a specificity of 99.2 (95% CI: 96.9-99.9) and 99.6 (95% CI: 97.5-99.9), respectively (Table 2).

**Table 2:** Specificity of sVNT

| Category | Total (n) | 20% Cut-off |  |  | 30% Cut-off |  |  |
| --- | --- | --- | --- | --- | --- | --- | --- |
|  |  | Positive (%) | Negative (%) | Specificity (95% CI) | Positive (%) | Negative (%) | Specificity (95% CI) |
| Pre-pandemic adult patients | 21 | 0 (0) | 21 (100) | 100 (80.8-100) | 0 (0) | 21 (100) | 100 (80.8-100) |
| Pre-pandemic child patients | 50 | 0 (0) | 50 (100) | 100 (91.1-100) | 0 (0) | 50 (100) | 100 (91.1-100) |
| CMV | 10 | 0 (0) | 10 (100) | 100 (65.5-100) | 0 (0) | 10 (100) | 100 (65.5-100) |
| EBV | 10 | 0 (0) | 10 (100) | 100 (65.5-100) | 0 (0) | 10 (100) | 100 (65.5-100) |
| Suspected Pertussis | 16 | 0 (0) | 16 (100) | 100 (75.9-100) | 0 (0) | 16 (100) | 100 (75.9-100) |
| Other Resp. Diseases | 52 | 1 (1.9) | 51 (98.1) | 98.1 (88.4-99.9) | 0 (0) | 52 (100) | 100 (91.4-100) |
| Blood Donors | 100 | 1 (1.0) | 99 (99.0) | 99.0 (93.8-99.9) | 1 (1.0) | 99 (99.0) | 99.0 (93.8-99.9) |
| Total | 259 | 2 (0.8) | 257 (99.2) | 99.2 (96.9-99.9) | 1 (0.4) | 257 (99.6) | 99.6 (97.5-99.9) |

CMV, cytomegalovirus; EBV, Epstein-Barr virus; ILI, influenza like illness; 95% CI, 95% confidence interval

**Figure 1:**
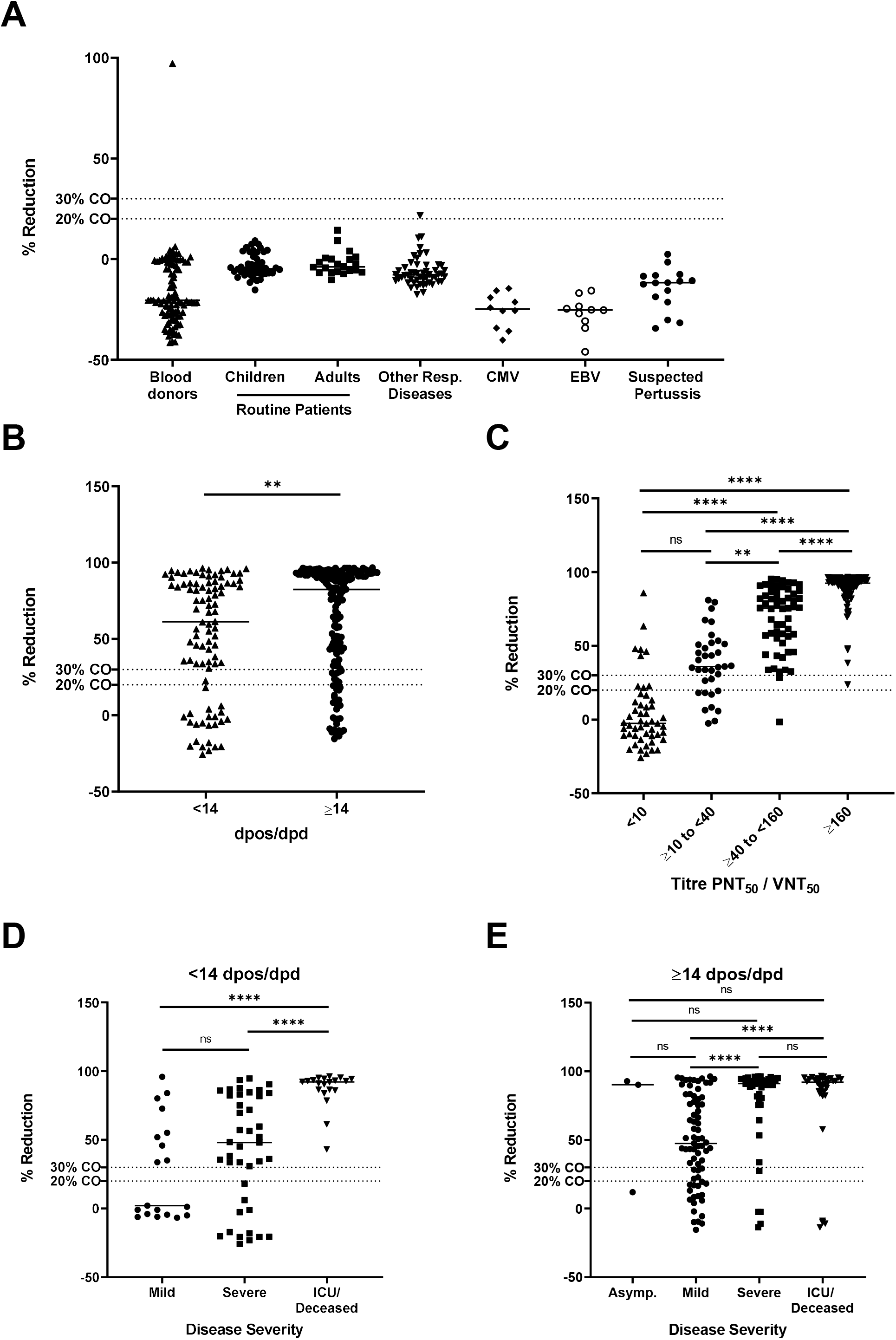
%reduction (inhibition of RBD-ACE2 binding) of samples of the specificity panel). %reduction of PCR-confirmed COVID-19 patient samples stratified **(A)** by days post onset of symptoms (dpos) or days post diagnosis (dpd) (**B**), by results of PNT50/VNT50 titre (**C**) and by disease severity (**D** and **E**). Dashed line indicates 20% or 30% cut-off (CO).

Among the 269 sera of confirmed COVID-19 patients in the sensitivity panel, 216 sera tested positive in the sVNT resulting in an overall clinical sensitivity of 80.3 (95%CI: 74.9-84.8). In sera sampled before 14 dpos/dpd 69/92 samples blocked RBD-ACE2 interaction (clinical sensitivity 75.0 (95%CI: 64.7-83.2) while 147/177 sera sampled at or after 14 dpos/dpd tested positive, resulting in an increased clinical sensitivity of 83.1 (95%CI: 76.5-88.1). In addition, median %reduction significantly increased between < 14 and ≥14 dpos/dpd from 61.2% to 82.5% (Mann Whitney test, p=0.0019) (Figure 1B, 2A and Table 3). Furthermore, we analysed the relationship between the severity of COVID-19 disease and the clinical sensitivity of the sVNT as stronger immune responses were observed in severe vs mild cases^14,15^. We observed that the assay sensitivity increased with an increased disease severity, higher in hospitalised patients vs outpatients, regardless of the time of sampling. During the acute phase (< 14dpos/dpd) of the infection, the clinical sensitivity in mild cases was 47.4% and increased to 70.7% and 100% in severe and ICU/deceased patients, respectively (Figure 1D and Table 3). A similar observation was made in samples collected ≥14 dpos/dpd, where sensitivity increased from 75.3% in mild cases to 91.3% and 88.2% in severe and ICU/deceased cases, respectively (Figure 1E and Table 3). The median %reduction in the sVNT increased with disease severity, but this increase was only significant for mild vs severe and mild vs ICU/deceased cases both before and after 14dpos/dpd (Kruskal-Wallis test) (Figure 1D and E). When considering the alternative cut-off of 30% reduction to determine positivity in the sVNT, the clinical sensitivity of the assay decreased by 0-9%, but the overall picture remained unchanged (Table 3).

**Table 3:** Sensitivity of sVNT

| Category | Total | 20% Cut-off |  |  | 30% Cut-off |  |  |
| --- | --- | --- | --- | --- | --- | --- | --- |
|  |  | Positive (%) | Negative (%) | Sensitivity % (95% CI) | Positive (%) | Negative (%) | Sensitivity % (95% CI) |
| All | 269 | 216 (80.3) | 53 (19.7) | 80.3 (74.9-84.8) | 207 (77.0) | 62 (23.0) | 77.0 (71.4-81.8) |
| dpos/dpd |  |  |  |  |  |  |  |
| <14 days | 92 | 69 (75.0) | 23 (25.0) | 75.0 (64.7-83.2) | 68 (73.9) | 24 (26.1) | 73.9 (63.5-82.3) |
| ≥14 days | 177 | 147 (83.1) | 30 (16.9) | 83.1 (76.5-88.1) | 139 (78.5) | 38 (21.5) | 78.5 (71.6-84.2) |
| P-NT50/VNT50 |  |  |  |  |  |  |  |
| <10 | 50 | 9 (18.0) | 41 (82.0) | 18.0 (9.0-31.9) | 6 (12.0) | 44 (88.0) | 12.0 (5.0-25.0) |
| ≥10 to <40 | 35 | 26 (74.3) | 9 (25.7) | 74.3 (56.4-86.9) | 24 (68.6) | 11 (31.4) | 68.6 (50.6-82.6) |
| ≥40 to <160 | 57 | 56 (98.2) | 1 (1.8) | 98.2 (89.4-99.9) | 55 (96.3) | 2 (3.7) | 96.3 (86.2-99.4) |
| ≥160 | 116 | 116 (100) | 0 (0) | 100 (96.0-100) | 115 (99.1) | 1 (0.9) | 99.1 (94.6-100) |
| ND | 11 | 9 (81.8) | 2 (18.2) | 81.8 (47.8-96.8) | 7 (63.6) | 4 (36.4) | 63.6 (31.6-87.6) |
| Severity <14 dpos/dpd |  |  |  |  |  |  |  |
| Mild | 19 | 9 (47.4) | 10 (52.6) | 47.4 (25.2-70.5) | 9 (47.4) | 10 (52.6) | 47.4 (25.2-70.5) |
| Severe | 41 | 29 (70.7) | 12 (29.3) | 70.7 (54.3-83.4) | 29 (70.7) | 12 (29.3) | 70.7 (54.3-83.4) |
| Deceased | 20 | 20 (100) | 0 (0) | 100 (80.0-100) | 20 (100) | 0 (0) | 100 (80.0-100) |
| unknown | 12 | 11 (91.7) | 1 (8.3) | 91.7 (59.8-99.6) | 10 (83.3) | 2 (16.7) | 83.3 (50.9-97.1) |
| Severity ≥14 dpos/dpd |  |  |  |  |  |  |  |
| Asymptomatic | 3 | 2 (66.7) | 1 (33.3) | 66.7 (12.5-98.2) | 2 (66.7) | 1 (33.3) | 66.7 (12.5-98.2) |
| Mild | 73 | 55 (75.3) | 18 (24.7) | 75.3 (63.6-84.4) | 50 (68.5) | 23 (31.5) | 68.5 (56.4-78.6) |
| Severe | 46 | 42 (91.3) | 4 (8.7) | 91.3 (78.3-97.2) | 41 (89.1) | 5 (10.9) | 89.1 (75.6-95.9) |
| ICU/Deceased | 34 | 30 (88.2) | 4 (11.8) | 88.2 (71.6-96.2) | 30 (88.2) | 4 (11.8) | 88.2 (71.6-96.2) |
| unknown | 21 | 18 (85.7) | 3 (14.3) | 85.7 (62.6-96.2) | 16 (76.2) | 5 (23.8) | 76.2 (52.5-90.9) |

DPOS, days post onset of symptoms; DPD days post diagnosis; P-NT50 pseudovirus neutralization test 50% inhibition titer; VNT50 virus neutralization test 50% inhibition titer; ND, not determined; 95% CI, 95% confidence interval

As the novel assay is meant to be implemented as a surrogate assay for functional cell-based serological methods that measure SARS-CoV-2 neutralising capacity of antibodies, we compared the performance of the sVNT with the conventional VNT_50_ and PNT_50_ assays. The sVNT detected blocking of ACE2 binding activity in nine of 50 sera of confirmed COVID-19 patients that did not show SARS-CoV-2 neutralising activity in the cell-based assays (Table 3). Conversely, only 26/35 sera with a titre in the range of ≥10 to <40 as well as 56/57 sera with a titre in the range of ≥40 to <160 in the cell-based neutralisation assays showed blocking activity in the sVNT. This results in an analytical sensitivity of 74.3 (95% CI: 56.4-86.9) and 98.2 (95% CI: 89.4-99.9) respectively when compared to conventional in-house neutralisation assays. Only sera (n=116) with a titre of ≥160 in the PNT_50_/VNT_50_ were always positive in the sVNT. The median %reduction in the four titre subgroups significantly increased with rising PNT50/VNT50 titres from −2.6 (< 10) to 36.0 (≥10 to < 40), 75.6 (≥40 to < 160) and 92.5 (≥160) (Kruskal-Wallis test) (Figure 1C). To further investigate the correlation of PNT_50_/VNT_50_ titres with %reduction in the sVNT, a linear regression analysis was performed using all sera for which an endpoint titre was available (n=154). We found a moderate correlation between PNT_50_ and sVNT %reduction (R^2^= 0.4937, p>0.0001) (Figure 2B) and a slightly better correlation for VNT50 titres (R^2^ = 0.6548, p>0.0001) (Figure 2C).

**Figure 2:**
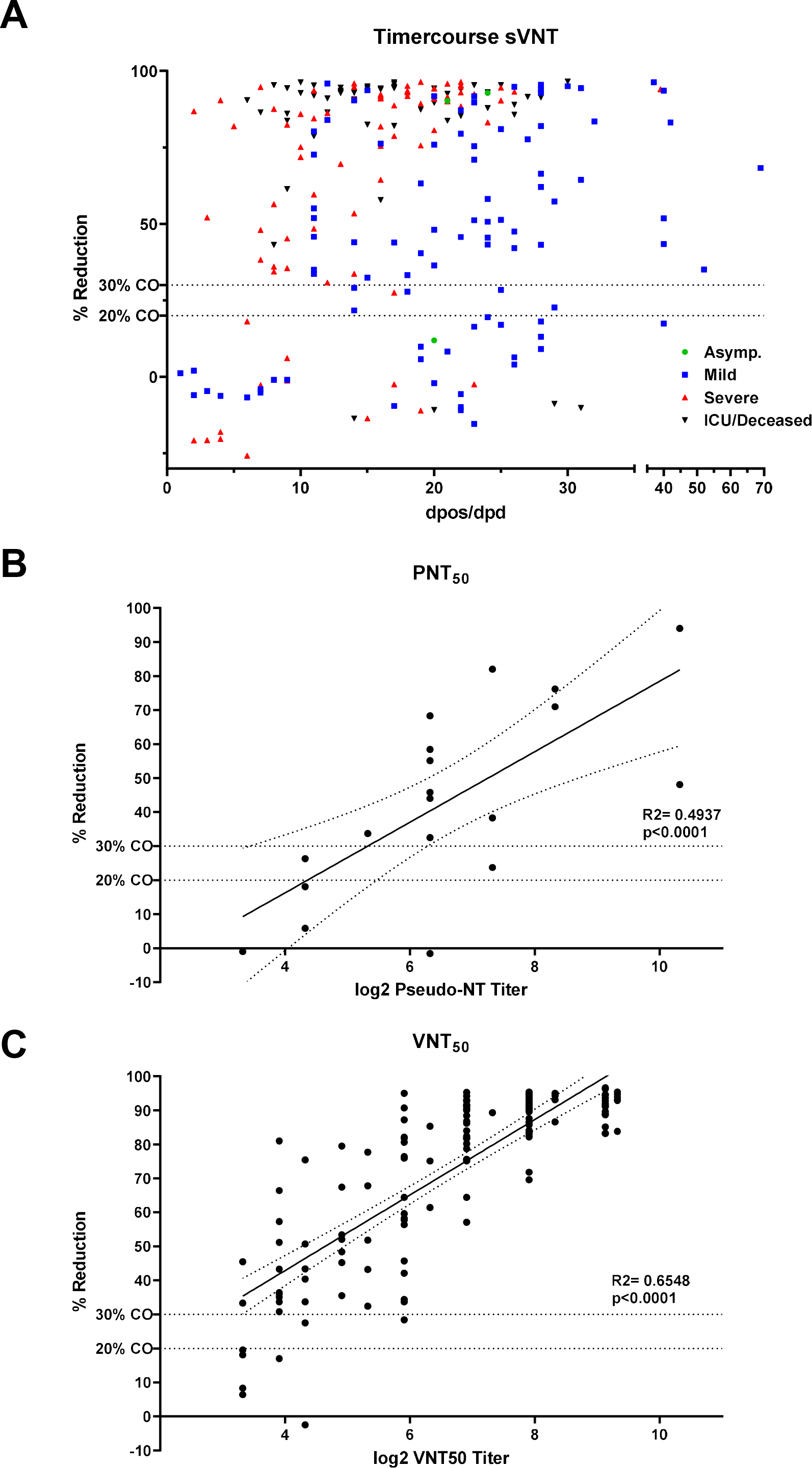
%reduction (inhibition of RBD-ACE2 binding) of COVID-19 patient samples by dpos/dpd and disease severity (**A**). Linear correlation of PNT_50_ (**B**) and VNT_50_ (**C**) endpoint titres with %reduction of RBD-ACE2 binding. Dashed line indicates 20% or 30% cut-off (CO).

We also compared overall clinical sensitivity and specificity of the assay between the two centres where the study was conducted (UNIGE and RIVM). Specificity was comparable with only minor differences between the two laboratories. In contrast, clinical sensitivity was markedly lower at UNIGE compared to samples analysed at RIVM (Table 4). However, samples from late time points dpos/dpd where underrepresented in the UNIGE sample set, giving a potential explanation for this discrepancy.

**Table 4:** Comparison of sVNT between sites

|  | UNIGE |  | RIVM |  |
| --- | --- | --- | --- | --- |
|  | 20% Cut-off | 30% Cut-off | 20% Cut-off | 30% Cut-off |
| Overall Specificity (95% CI) | 100 (93.6-100) | 100 (93.6-100) | 98.9 (95.8-99.8) | 99.5 (96.6-99.9) |
| Overall Sensitivity (95% CI) | 70.9 (56.9-81.9) | 63.6 (49.5-75.8) | 82.7 (76.8-87.4) | 80.4 (74.3-85.3) |

95% CI, 95% confidence interval

Last, we investigated the interassay variance of the sVNT by testing a representative subset of samples at least five times at different days. Similar coefficients of variation (%CV) were found in both laboratories. We observed a very low %CV in samples with a mean %reduction > 90%, ranging from 0.22 to 4.63 %CV. However, the %CV increased to 3.55-13.73 in samples with a mean %reduction between 50-60% and to 10.39-20.39 in samples with a mean %reduction around 30%. Samples with a mean %reduction < 11% gave a high %CV between 67.33-216.38 (Table 5).

**Table 5:**
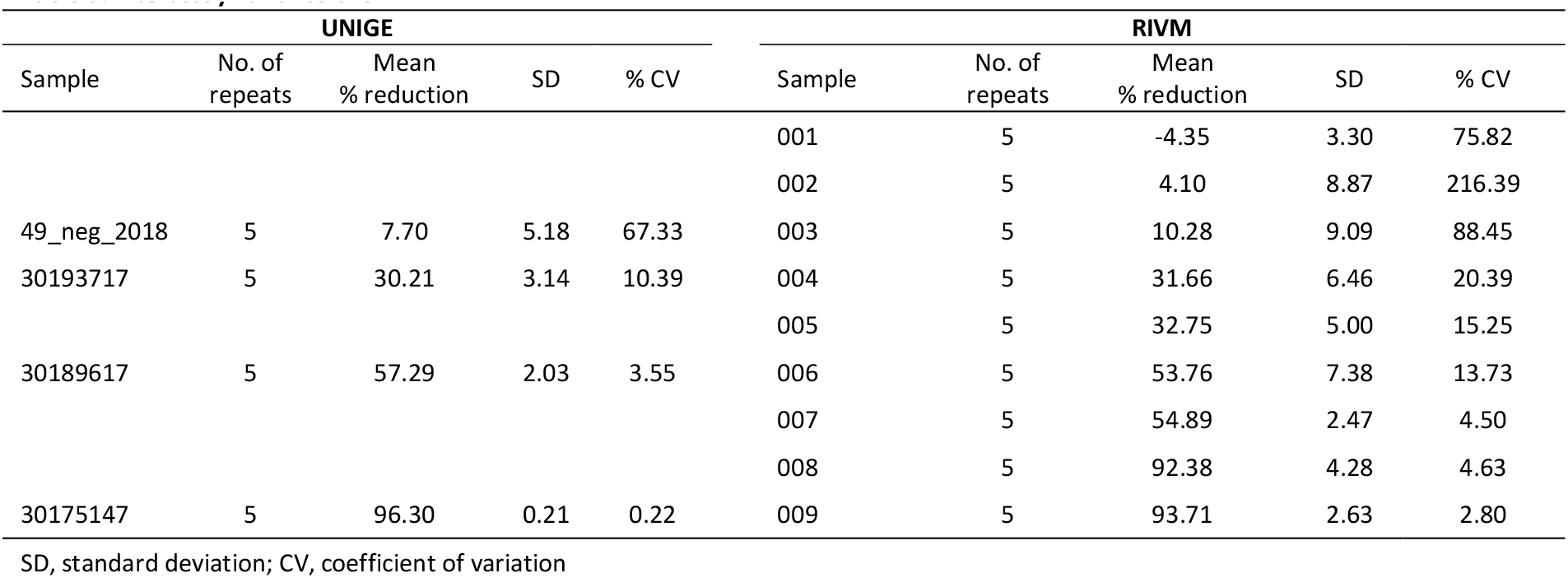
Interassay variance of sVNT

## Discussion

The importance of serology in clinical and public health management of SARS-CoV-2 is reflected in the huge amount of immune-assays that are currently being developed or have been released on the diagnostic market^1^. However, a vast majority of these tests are simply measuring qualitatively or semi-quantitatively the presence of IgG, IgM and/or IgA but do not address the functionality of the antibody response elicited by a SARS-CoV-2 infection. Functional assays like virus neutralization tests are essential to address specific questions related to protective immunity after vaccination or natural infection. However, these type of assays are operated based on in-house protocols and lack standardization across laboratories^16–18^. We performed an independent two-centre clinical evaluation of the GenScript cPass™ test, which is based on the principle of an inhibition ELISA, to assess its value for routine diagnostics and as surrogate functional assay to measure neutralizing capability in SARS-CoV-2 elicited antibody responses.

We observed a high specificity of > 99% and an overall clinical sensitivity of 83% in samples taken ≥14 dpos. Within this group of confirmed COVID-19 patients, the clinical sensitivity was 75% in mild and 91% in hospitalized patients. Deeks and colleagues performed a Cochrane assessment of 54 studies using commercial immune-assays^5^. They concluded that in confirmed COVID-19 patients sampled in the periods 15-21 dpos and 22-25 dpos, the sensitivities for IgG were respectively 88.2% (95% CI 83.5 to 91.8) and 80.3% (95%CI 72.4 to 86.4). While direct comparison of performances of different serology assays is complicated by differences in test set-up (e.g. testing for all isotypes vs for IgG only, the use of different antigens, measuring antigen binding vs prevention of RBD-ACE2 binding) and differences in patient cohorts used, our results seem to indicate that the sVNT might be a more powerful tool for cohort and population studies than for individual diagnosis of past SARS-CoV-2 infection as the observed sensitivities are in the lower end of the range of average test performances observed by Deeks.

With respect to the sVNT as alternative for conventional virus culture-based assays measuring neutralizing activity, we observed that on the one hand the sVNT misses samples that have a low virus neutralization titre in these assays and on the other hand identifies samples as positive that were negative in a Gold Standard tests. A limitation of the sVNT is the restriction to RBD binding antibodies. It has been shown that SARS-CoV-2 infection does not only induce antibodies against the RBD or the S1 domain, but also against the S2 domain as well as against N^19^. While antibodies directed against the N protein are most likely non-neutralizing, antibodies directed against the N-terminal domain of S1 (outside of the RBD) have shown neutralizing potential^20^. In addition, for SARS-CoV-1 also antibodies directed at the S2 domain show neutralizing capabilities^21^. Although S2 domain-mediated neutralisation remains to be confirmed for SARS-CoV-2, assessing the neutralizing ability with the sVNT might indeed miss the presence of virus neutralizing capabilities directed outside of the RBD. Furthermore, all of the sera that were reactive in the sVNT but not in the VNT_50_/PNT_50_ were reactive in the Wantai total Ig ELISA that targets RBD as well (data not shown). This indicates that the activity measured in the sVNT is likely due to antibodies directed against the RBD but without a virus neutralising capacity that leads to at least 50% reduction of infected cells. Another explanation for such false positivity with respect to neutralizing capability might be the presence of anti-ACE2 autoantibodies as described for specific patient groups^22,23^. The observation of false positives and the observed lower sensitivity (74%) of the sVNT in comparison to conventional tests in samples with neutralizing antibody titres ≥10 and < 40, indicate that the sVNT cannot fully replace the Gold Standard immune-assays. Depending on the context of use, the lower sensitivity in the range of low virus neutralization titres might however be acceptable, e.g. for decision making on release of hospitalized patients with virus neutralization titres above a predetermined threshold from isolation units^24,25^. Nevertheless, as long as the correlates of protection for classical virus neutralization tests are not known, the use of the sVNT as functional assay to determine level of immunity is not warranted.

Besides the aforementioned advantages of the possibility for increased standardization across laboratories, the possibility for automatization, the technical simplicity and the reduced biosafety risk, the sVNT is isotype- and species-independent. Species-independent serology tools are important for research into the epidemiology and ecology of SARS-CoV-2, i.e. for the identification of natural reservoirs and spill-over hosts as well as the monitoring and prevention of human risks from sustained virus circulation in farm animals such as minks (refs). In contrast to virus culture-based assays, the sVNT is at most a semi-quantitative assay making it less valuable as functional assay in immunity studies.

In conclusion, the sVNT can be used as an additional assay to determine the immune status of COVID-19 infected of vaccinated individuals and cohort studies to confirm results of more routine immuno-assays like IgG, IgM and/or IgA ELISAs and CLIAs. The value of the sVNT as functional assay in patient management, biosafety management, vaccine and immunity studies needs to be assessed for the specific context of use.

## Data Availability

The data will be made available upon reasonable request.

## Acknowledgements

We would like to thank Romain Burquier (HUG, Geneva) for technical assistance and Barbara Lemaitre (HUG, Geneva) and Shefije Fejza (HUG, Geneva) for help with sample preparation. We are grateful to Jean-Luc Murk (Elizabeth-Tweesteden Ziekenhuis, Tilburg, the Netherlands), Afke Brandenburg (Izore, Leeuwarden, the Netherlands) and Bas Wintermans (Admiraal de Ruyter Ziekenhuis, Goes, the Netherlands), Josine van Beek and Kristin Kremer (RIVM) for provision of panel samples an Mariette Edema (RIVM) for support. This study was support by a grant from the Private Foundation of the Geneva University Hospital.

## Bibliography

1. Foundation for Innovative New Diagnostics. FIND evaluation update: SARS-CoV-2 immunoassays. FIND evaluation update: SARS-CoV-2 immunoassays https://www.finddx.org/covid-19/sarscov2-eval-immuno/ (2020).

2. Meyer, B. et al. Validation of a commercially available SARS-CoV-2 serological immunoassay. Clinical Microbiology and Infection S1198743X20303682 (2020) doi:10.1016/j.cmi.2020.06.024.

3. Lassaunière, R. et al. Evaluation of nine commercial SARS-CoV-2 immunoassays. medRxiv 2020.04.09.20056325 (2020) doi:10.1101/2020.04.09.20056325.

4. GeurtsvanKessel, C.H. et al. An evaluation of COVID-19 serological assays informs future diagnostics and exposure assessment. Nat Commun 11, 3436 (2020).

5. Deeks, J.J. et al. Antibody tests for identification of current and past infection with SARS-CoV-2. Cochrane Database of Systematic Reviews (2020) doi:10.1002/14651858.CD013652.

6. Barnes, C.O. et al. Structures of Human Antibodies Bound to SARS-CoV-2 Spike Reveal Common Epitopes and Recurrent Features of Antibodies. Cell 182, 828–842.e16(2020).

7. Tan, C.W. et al. A SARS-CoV-2 surrogate virus neutralization test (sVNT) based on antibody-mediated blockage of ACE2-spike (RBD) protein-protein interaction. https://www.researchsquare.com/article/rs-24574/v1 (2020) doi:10.21203/rs.3.rs-24574/v1.

8. Wrapp, D. et al. Cryo-EM structure of the 2019-nCoV spike in the prefusion conformation. Science eabb2507 (2020) doi:10.1126/science.abb2507.

9. Hoffmann, M. et al. SARS-CoV-2 Cell Entry Depends on ACE2 and TMPRSS2 and Is Blocked by a Clinically Proven Protease Inhibitor. Cell S0092867420302294 (2020) doi:10.1016/j.cell.2020.02.052.

10. van Beek, J. et al. Influenza-like Illness Incidence Is Not Reduced by Influenza Vaccination in a Cohort of Older Adults, Despite Effectively Reducing Laboratory-Confirmed Influenza Virus Infections. The Journal of Infectious Diseases 216, 415–424 (2017).

11. Berger Rentsch, M. & Zimmer, G. A vesicular stomatitis virus replicon-based bioassay for the rapid and sensitive determination of multi-species type I interferon. PLoS ONE 6, e25858 (2011).

12. Torriani, G. et al. Identification of Clotrimazole Derivatives as Specific Inhibitors of Arenavirus Fusion. J.Virol. 93, (2019).

13. Fukushi, S. et al. Vesicular stomatitis virus pseudotyped with severe acute respiratory syndrome coronavirus spike protein. J.Gen. Virol. 86, 2269–2274 (2005).

14. Rijkers, G. et al. Differences in Antibody Kinetics and Functionality Between Severe and Mild Severe Acute Respiratory Syndrome Coronavirus 2 Infections. The Journal of Infectious Diseases jiaa463 (2020) doi:10.1093/infdis/jiaa463.

15. Cervia, C. et al. Systemic and mucosal antibody secretion specific to SARS-CoV-2 during mild versus severe COVID-19. http://biorxiv.org/lookup/doi/10.1101/2020.05.21.108308 (2020) doi:10.1101/2020.05.21.108308.

16. Muruato, A.E. et al. A high-throughput neutralizing antibody assay for COVID-19 diagnosis and vaccine evaluation. Nat Commun 11, 4059 (2020).

17. Zettl, F. et al. Rapid Quantification of SARS-CoV-2-Neutralizing Antibodies Using Propagation-Defective Vesicular Stomatitis Virus Pseudotypes. Vaccines 8, 386 (2020).

18. Wölfel, R. et al. Virological assessment of hospitalized patients with COVID-2019. Nature 581, 465–469 (2020).

19. Grzelak, L. et al. A comparison of four serological assays for detecting anti–SARS-CoV-2 antibodies in human serum samples from different populations. Sci. Transl. Med. 12, eabc3103 (2020).

20. Chi, X. et al. A neutralizing human antibody binds to the N-terminal domain of the Spike protein of SARS-CoV-2. Science 369, 650–655 (2020).

21. Duan, J. et al. A human SARS-CoV neutralizing antibody against epitope on S2 protein. Biochem. Biophys. Res. Commun. 333, 186–193 (2005).

22. Takahashi, Y., Haga, S., Ishizaka, Y. & Mimori, A. Autoantibodies to angiotensin-converting enzyme 2 in patients with connective tissue diseases. Arthritis Res Ther 12, R85 (2010).

23. McMillan, P. MB,BS,MRCP, & Uhal, B.D., PhD. COVID-19–A theory of autoimmunity to ACE-2. MOJ Immunology 7, 17–19 (2020).

24. Singanayagam, A. et al. Duration of infectiousness and correlation with RT-PCR cycle threshold values in cases of COVID-19, England, January to May 2020. Eurosurveillance 25, (2020).

25. van Kampen, J. J. A. et al. Shedding of infectious virus in hospitalized patients with coronavirus disease-2019 (COVID-19): duration and key determinants. http://medrxiv.org/lookup/doi/10.1101/2020.06.08.20125310 (2020) doi:10.1101/2020.06.08.20125310.

